# Early Sport Specialization Impact on Rates of Injury in Collegiate and Professional Sport Participation: A Systematic Review and Meta-analysis

**DOI:** 10.1101/2023.07.31.23293438

**Authors:** Bahman Adlou, Wendi Weimar, Christopher Wilburn, Alan Wilson

## Abstract

Early sport specialization (ESS) is common among adolescent athletes due to desire of reaching elite status; however, the long-term impact of ESS on sports injury (SI) rates is not fully understood. We performed a systematic review and meta-analysis of the literature to evaluate the effect of ESS on SI rates during elite sport participation. Search included PubMed, Web of Science, SPORTDiscus, EMBASE, Cochrane, CINAHL, ERIC, and Medline databases. Preferred Reporting Items for Systematic Reviews and MetaAnalyses (PRISMA) guidelines were followed to identify published peer-reviewed articles from inception to 23 March 2023. Eligible studies reported numbers of elite athletes in collegiate, national, and professional teams in ESS and their counterpart category, as well as the portion of each group with at least one SI. Studies on concussion or non-contact injuries, or without explicit injury reports during elite sprot participation were excluded. Five articles were included in the review; 3 reported SIs on collegiate and 2 reported on professional athletes. A total of 3087 athletes were included (ESS = 45%). The overall effect was not significant and demonstrated that ESS athletes had a lower odd (OR=0.7) of a SI during elite sport participation. Articles with clear reports are scarce, and thus limit the impact of the findings in this study. High heterogeneity was evident in this analysis, largely due to lack of standardized terminology, evaluation, and data representation. Prospective studies that consider diverse athlete population of the same sport are warranted.

## Introduction

Participating in organized sports (e.g., professional and school teams, traveling teams, city clubs, or off-season camps sports) has become increasingly common among middle- and high school aged adolescents. According to the National Federation of State High School Associations, the number of adolescent athletes in the United States (U.S.) grew by nearly one million over the last decade, reaching to approximately 7.8 million adolescent athletes[1]. For many of these young athletes, there is pressure from their peers, parents, and coaches to focus on one sport[2–5]. This sport participation pattern is commonly referred to as *early sports specialization* (ESS) in the literature[6,7], which is defined as year-round single-sport participation at the exclusion of other sports from a young age[8,9]. Although sport participation comes with numerous benefits, such as improved physical fitness and social development especially for adolescents, ESS reduces unstructured and free play [10], with evidence indicating that it increases rates of overuse injuries, burnouts, and even withdrawal from the sport[11,12].

The increasing pressure on adolescent athletes for ESS in the U.S. is fueled in part by the popular belief that ESS leads to increased success with scholarship attainment at collegiate level, reaching elite athletic status (e.g., national team), receiving endorsement and sponsorships, or signing professional sports contracts[2–5]. However, there is evidence that suggests this may not be the case[13,14], and the potential negative consequences of ESS including increased risk of injury outweigh the benefits[15]. This concern has prompted involvement of governing bodies and sports science and medicine professionals to educate the public on the risk-benefit ratio of ESS among adolescent athletes.

The American Orthopaedic Society for Sports Medicine (AOSSM) published a consensus statement in 2016, which recommended that young athletes should delay specialization until at least late adolescence and should participate in multiple sports throughout childhood [10]. The American Medical Society for Sports Medicine (AMSSM) Youth Early Sport Specialization Summit in 2018 expanded upon the AOSSM statement to define a research agenda for youth sport specialization in the U.S. [16], which highlighted the need for further research to better understand the risks and benefits of ESS. A recent comprehensive review found athletes with high sport specialization at an increased risk of sustaining an overuse injury compared with multisport and low sport specialized athletes, and concluded that ESS is associated with an increased risk of overuse and musculoskeletal injuries[17].

Various studies have also investigated the long-term effects of ESS on injury rates, such as in collegiate leagues. In a study on collegiate athletes, Post et al. (2017) found that those who specialized in their collegiate sport before the age of 12 had a significantly higher risk of lower extremity injuries compared to those who did not specialize early[18]. In another study, the authors analyzed sports specialization in NCAA Division I athletes and found that single-sport-specialized athletes were at a greater risk of injury than those who participated in multiple sports before the age of 12[17]. These findings suggest that ESS could contribute to the increased risk of sports injuries in elite sport participation, including the collegiate, national, or international leagues.

Overall, the literature suggests that ESS may lead to increased risk of sport injuries and does not necessarily increase the chances of achieving elite athletic status. We hypothesized that elite athletes who specialize in a single-sport early (ESS) present with a higher rate of injuries in their elite sport participation (i.e., collegiate, professional, national/international level) than those who did not specialize early. A comprehensive evaluation of the association between ESS and injury rates among athletes will aid the informed group decision-making by the adolescent athlete and their parents and coaches before committing to a single-sport participation at a young age. However, there is no quantitative analysis of the peer-reviewed data to show if individuals who specialized in their sport early sustained injuries at a greater rate than those who were not early specializers. Therefore, in this systematic review and meta-analysis, we quantitively evaluate the impact of ESS on injury rates in a sample of collegiate, national, and professional athletes.

## Methods

### Search Strategy and selection criteria

A systematic search was performed in accordance with updated Preferred Reporting Items for Systematic Reviews and Meta-Analysis (PRISMA) guidelines[19]. The electronic search was executed by two independent reviewers (BA, WW) through the following databases: Web of Science, Cochrane, PubMed, EMBASE, and ESBSCO-hosted registries, including SPORTDiscus, Cumulative Index to Nursing and Allied Health Literature (CINAHL), Educational Resources Information Center (ERIC), and Medline from inception until 23 March 2023. The search query was prepared by the lead author (BA) using Boolean operators in the following format: ((“sport specialization” OR “sport specialisation” OR year-round OR single?sport OR multi?sport OR sport?sampling) AND (young OR youth OR pediatric OR high?school OR child* OR adolescen*) AND (injur* OR sport?injury OR injury risk) AND (elite OR pro OR professional OR collegiate OR college OR league OR national OR Olympic)).

Duplication and the eligibility criteria, including language and article type, were automatically filtered. All remaining article titles and abstracts were independently screened for eligibility by two reviewers (BA, WW). Full texts of all potentially eligible articles were retrieved, or requested where necessary, for independent eligibility criteria screening by each reviewer. Articles that met the following eligibility criteria were included in this review and meta-analysis:

### Inclusion Criteria

- Articles published in English or Farsi languages.
- Included athlete non-contact musculoskeletal injury quantities during elite sport participation.
- Reports of any injury types that resulted in permanent or temporary (>3-day) cease of sport participation.
- Clear statement of *elite sport status*, defined by the athlete participating in any of the following leagues worldwide: collegiate, professional leagues, national leagues, and Olympics or other world-stage competitions. Under-14 years old worldwide competition was not considered as elite sport participation status.
- Studies must include early sport specialization (ESS) definition matching one of the following throughout adolescence sport participating before elite status: participating in one sport 8+ month year-round; participating in one sport at the exclusion of others (i.e., single-sport participation), or used 3-point sport specializing criteria [20]. Non-early sport specialization (non-ESS) were those groups in opposition of the above statements or defined as “multisport athletes” during adolescence before elite status by the authors.
- Early sport specialization age cut-off from 12 years old and older for both male and female athletes.

### Exclusion Criteria

- Concussion and direct-contact injuries
- Case studies, abstracts, and conference proceedings
- Combination of injury histories (rates or numbers) from non-elite sport participation for elite athletes and injury rates from elite sport participation (e.g., included injury quantities during youth non-elite sports for the currently elite athletes).
- Sport specialization 3-point criteria [20] without comparable non-specialized group.

After full-text review independently by authors (BA, WW), studies in agreement by both reviewers were included for analysis, and a third reviewer (CW) screened the studies that were marked eligible by only one of the reviewers. Additionally, the third reviewer served as the tiebreaker for inclusion of studies where necessary. All three reviewers (BA, WW, CW) collectively screened all other articles from citations. To be included, studies had to present a clear total number of athletes in each group, ESS versus late specializers (non-ESS), and the number of athletes from each group who reported at least one injury in their elite sport participation. A final list of included articles was aggregated (Microsoft Excel, v. 2303) through consensus for data extraction.

### Data extraction

Data extraction was independently done by two reviewers (BA, WW). A systematic extraction form (Microsoft Excel, v. 2303) was used for each article in full review to collect the following data along with the article info: (1) sample descriptions for each participant category (e.g., sample size and mean age across categories); (2) stage and type of elite sport participation (e.g., collegiate basketball or U.S. National Football League); and (3) study method (e.g., retrospective survey or descriptive epidemiology study). Data categories included the number of athletes in each of the following four categories: early sport specialized, number of early sport specializers with injuries, late sport specialized, and the number of late sport specializers with injuries. Reviewed article data in table format were transcribed manually, and data in figure plot format were transcribed using the updated version of a dedicated R Programming package [21]. Extracted data were compared and a final dataset was established for data analysis through consensus by all three reviewers (BA, WW, CW).

### Primary outcome

The primary outcome was to evaluate the impact of early sport specialization on long term rates of non-contact, sport injuries.

### Data analysis

Natural logarithm of odds ratio (logOR) was used to calculate the effect size for each study, along with the variance as following:

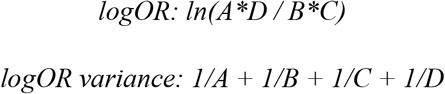

where A and C are the numbers of uninjured and injured ESS athletes, respectively, and B and D are the numbers of uninjured and injured non-ESS athletes, respectively. A positive logOR (or an Odds ratio of less than 1) indicates that more of those who specialized in sports early (ESS) suffered from injuries in their elite sports participation compared to non-ESS. This model accounts for both within-study variability and between-study heterogeneity.

The meta-analysis was performed using a weighted random-effects model with the restricted maximum likelihood (REML) estimator[22]. The estimated tau-squared (τ^2^) value was used as an indicator of the extent of heterogeneity across the included studies. Additionally, the I^2^ statistic was computed to assess the proportion of total variability attributable to heterogeneity. Q statistic was evaluated to assess the presence of heterogeneity, and its associated p-value was used to determine if the observed heterogeneity is statistically significant (i.e., not likely due to chance alone). The model results were reported as the estimated effect size (logOR), its standard error (SE), z-value, and p-value. The 95% confidence interval (CI) for the effect size was calculated to provide a range of plausible values. All statistical analyses and visualization were executed using RStudio (Posit Software. PBC, Build 386) via the *metafor* package (v. 4.2-0)[23].

### Sensitivity analysis and Risk of Bias

To assess publication bias, Egger’s test was performed[23]. Nonparametric (rank-based) trim-and-fill method by Duval and Tweedie [24] was used to estimate the number of studies missing from the analysis due to suppression of the most extreme results on one side of the funnel plot (funnel plot asymmetry). To assess the robustness of the results, a leave-one-out sensitivity was performed. By repeating the meta-analysis multiple times and leaving out one study each time, the influence of single study on the overall results was evaluated. Various influence measures can be used to assess the influence of individual studies on the overall results of the meta-analysis and to check the robustness of the meta-analysis, the influence test was performed.

## Result

### Search Results

The search strategy resulted in 526 total studies. Seven additional studies were identified through other sources (Figure 1).

**Figure 1.**
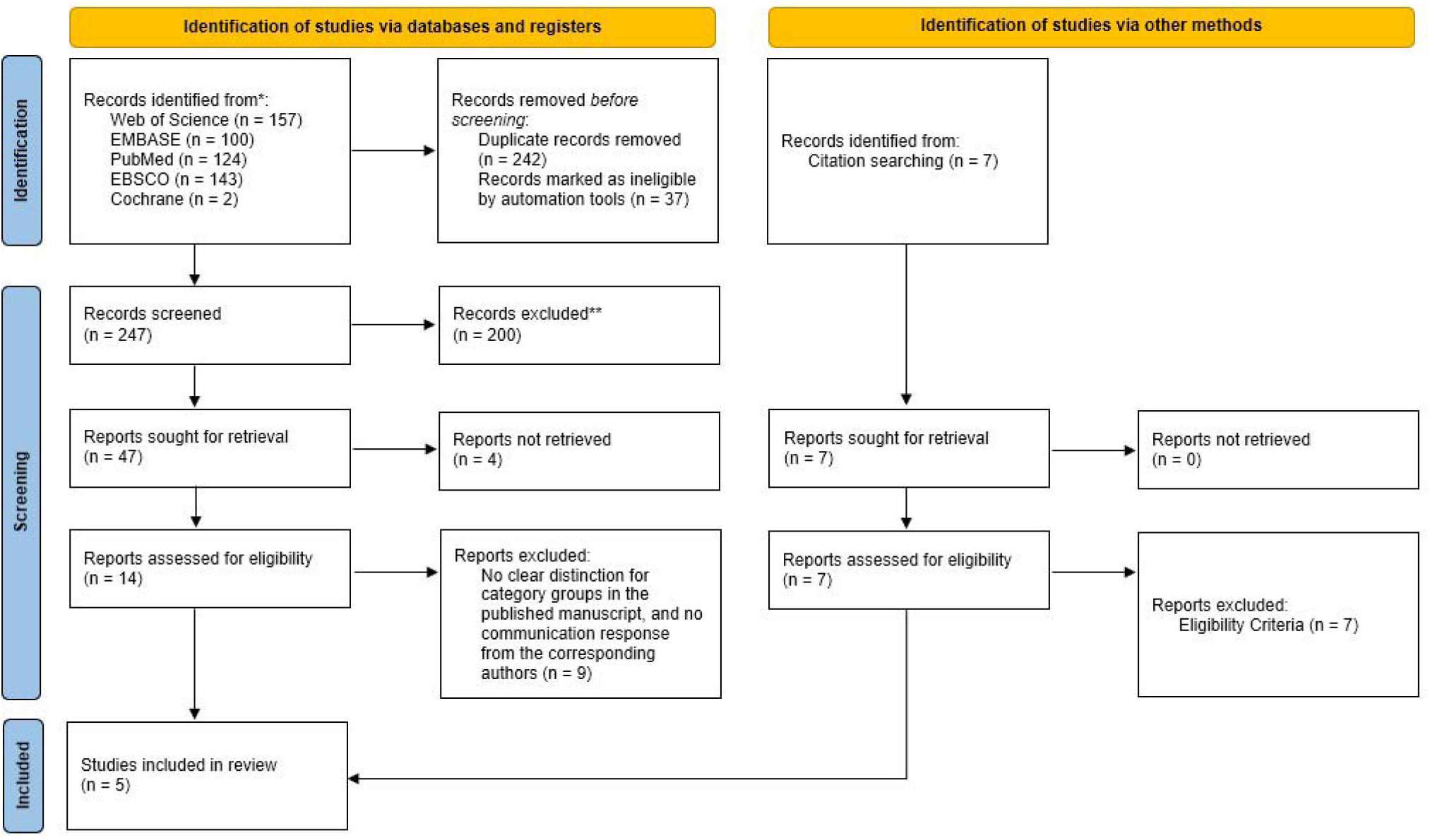
Flowchart of study selection. Flowchart adapted from the PRISMA 2020 statement.

Following removal of articles for duplication, article type, and language criteria, a total of 247 potentially eligible studies remained for title and abstract screening, from which two hundred studies were removed before full-text screening per eligibility criteria. Of the remaining forty-seven, 13 studies were agreed upon by all three reviewers for data extraction. Where possible, corresponding authors to each study were contacted and followed-up with to seek clarification or gain access to the dataset used in their studies. Of the 13 studies[5,25–36], five studies were included in the systematic review and meta-analysis due eligibility criteria or accessibility to data for analysis[5,25,29,30,34].

### Study characteristics

A total of 3087 athletes from five studies were included in this study for analysis. Forty-five percent (1396) of the athletes were identified as early sport specializers. Four studies reported on the sex of the athlete, which aggregated to 1989 males and 915 female athletes (Table 1).

**Table 1.** Summary Table of the athletes from the included studies; N = number of participants; ESS = early sport specialized; non-ESS = non-early sport specialized.

| Study | Elite | Total | Male | Female | ESS N | ESS | non- | non-ESS |
| --- | --- | --- | --- | --- | --- | --- | --- | --- |

|  | *status* | *Athletes N* | *Athletes N* | *Athletes N* |  | *Injured N* | *ESS N* | *Injured N* |
| --- | --- | --- | --- | --- | --- | --- | --- | --- |
| *Ahlquist et al. (2020)* | NCAA DI | 183 | 61 | 141 | 99 | 45 | 84 | 32 |
| *Rugg et al. (2021)* | NCAA DI | 1550 | 1006 | 544 | 280 | 84 | 1270 | 452 |
| *Confino et al. (2019)* | MLB | 746 | 746 | 0 | 506 | 216 | 240 | 110 |
| *Rugg et al. (2018)* | NBA | 237 | 237 | 0 | 201 | 86 | 36 | 9 |
| *Sweeney et al. (2021)* | NCAA DI | 371 | 0 | 371 | 310 | 269 | 61 | 41 |
| *Total* |  | 3087 | 2050 | 1056 | 1396 | 700 | 1691 | 644 |

The average age for early sport specialization was 14.3 years old with a standard deviation of 0.5 for the four studies that reported clear ESS age cut-off[5,25,30,34]. None of the studies were clinical trials or prospective studies. Three of the included studies reported on National Collegiate American Association (NCAA) Division I sport participation[5,25,30]; one reported on professional baseball sport participation (Major League Baseball in the U.S., or MLB) [29]; and the remaining study reported on professional men’s basketball (National Basketball Association in the U.S., or NBA) sport participation[34]. No further descriptive statistics or sub-analysis was possible due to inconsistencies in reporting the athlete characteristics in the included studies.

### Meta-analysis

A random-effects model was fitted to the five studies to estimate the overall effect of sports specialization on the odds of suffering from injury using the REML method. The estimated overall log odds ratio (logOR) is -0.314 with a standard error of 0.272 and the 95% confidence interval (CI) of -0.848, 0.22 (Figure 2).

**Figure 2.**
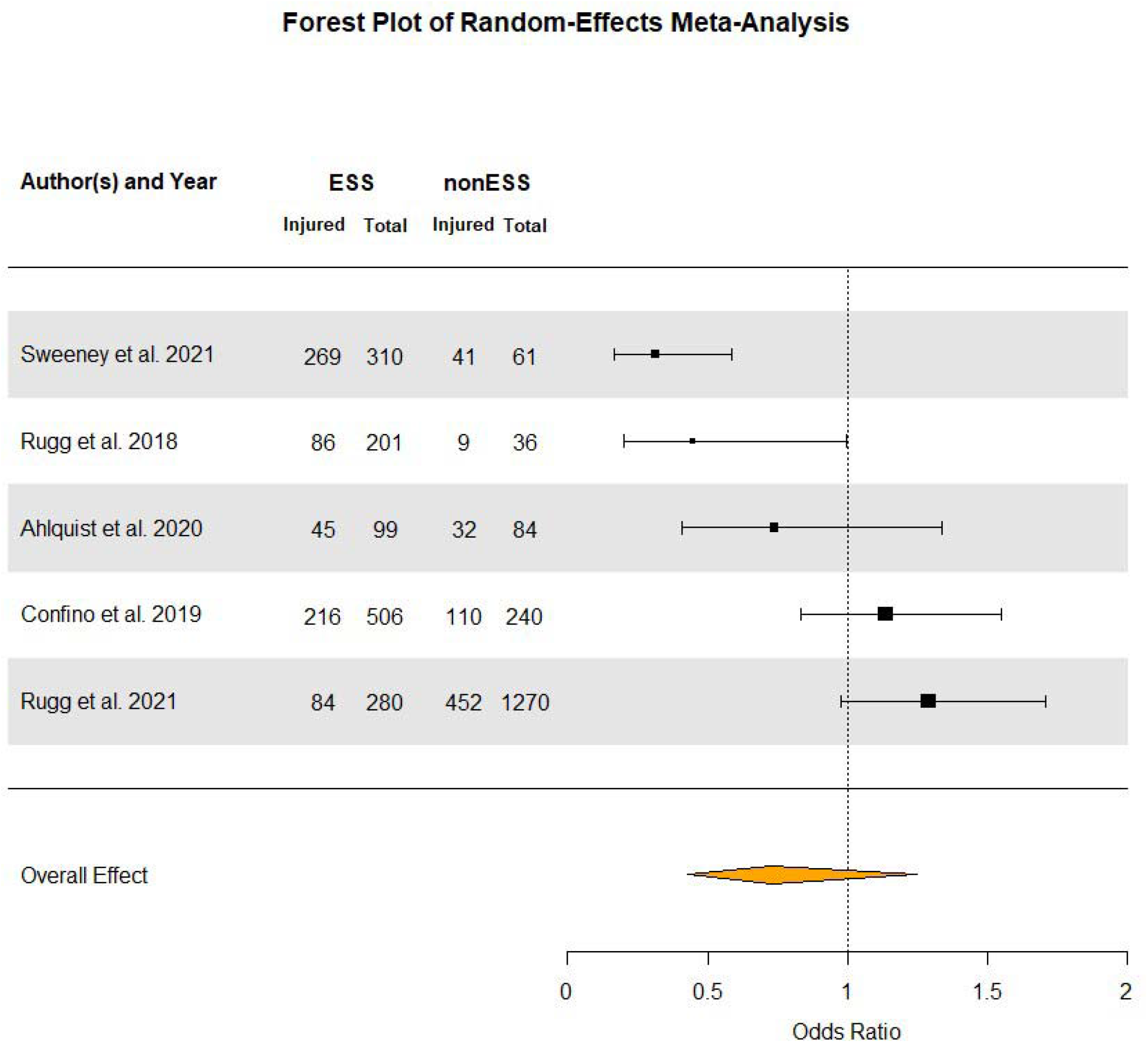
Forest plot of a random-effects meta-analysis of 5 studies examining the effect of early sports specialization on the odds ratio of non-contact injuries among elite athletes. The plot displays the individual study estimates and their corresponding confidence intervals, as well as the overall effect estimate. An overall effect of <1 indicates that those who specialize in their elite sport earlier had a lower odd (0.73) of being injured (as reported) compared to those who did not specialize in their sport early. However, the results are not statistically significant. ESS = total number of athletes in the early sport specialized group; ESS_injury = number of ESS athletes who reported to have at least 1 injury during elite sport participation; nonESS = total number of athletes in the non-early sport specialized group; nonESS_injury = number of nonESS athletes who reported to have at least 1 injury during elite sport participation.

To facilitate the interpretation of the results, logOR and its confidence interval was back transformed to odds ratios by taking the exponents such as following:

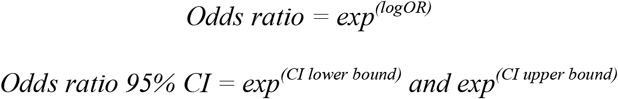

Odds ratio (OR) and its 95% CI associated with the findings are 0.731 and 0.428 to 1.246, respectively. However, this result was not statistically significant (p = 0.249). The estimated amount of total heterogeneity (τ^2^) is 0.297. The I^2^ statistic is 85.43%, suggesting that a large proportion of the total variability is due to heterogeneity between studies rather than sampling variability. The test for heterogeneity is significant (Q = 21.868, p < 0.001), indicating that the heterogeneity is statistically significant among the studies.

Funnel plot (Figure 3) graphical representations of the relationship between the effect size and the standard error of the effect size was executed.

**Figure 3.**
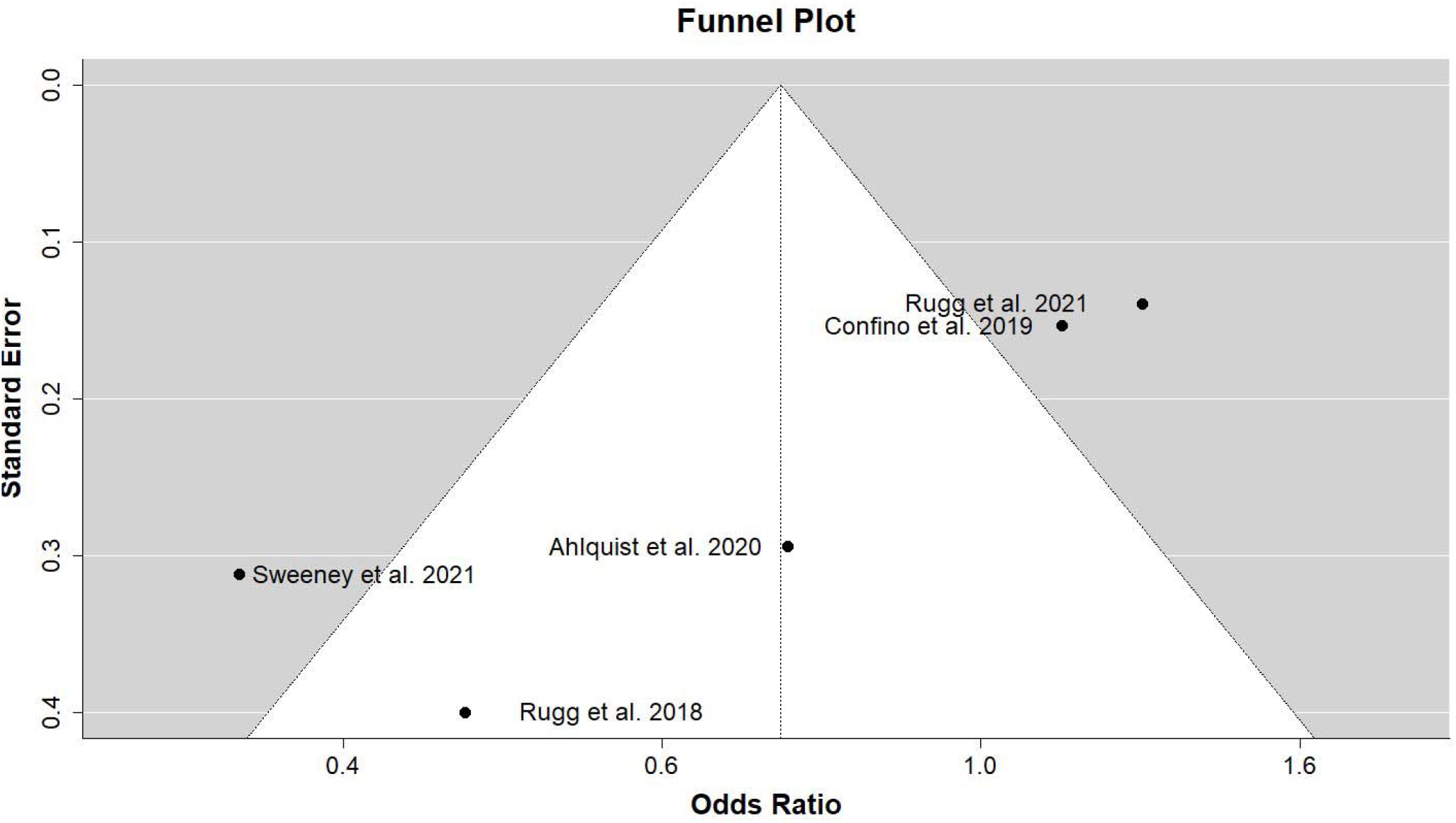
Funnel plot of the 5 studies included in the meta-analysis. The inverted funnel shape represents the expected distribution of studies in the absence of bias. Each point represents a study, with the horizontal axis showing the odds ratio and the vertical axis showing the standard error. The white area of the plot represents the values within the 95% confidence interval. Points falling outside of the confidence interval (represented by diagonal lines) may indicate potential publication bias or other small-study effects.

The trim-and-fill method did not impute any missing studies, and thus did not adjust the data. Egger’s test of funnel plot asymmetry output was significant (z-value = -4.244, p-value < 0.001), which indicates potential presence of publication bias. The leave-one-out sensitivity analysis showed that none of the individual studies had a large influence on the overall results of our meta-analysis. Removing any one study did not result in a statistically significant overall effect size estimate (i.e., all p-values > 0.05).

Influence analysis included studentized residuals, DFFITS values, Cook’s distance values, and covariance ratios. The results of this influence analysis showed that none of the individual studies had a large influence on the overall results of our meta-analysis according to any of these measures. In summary, our leave-one-out sensitivity and influence analysis suggest that our meta-analysis results are robust to the influence of individual studies.

## Discussion

The purpose of this systematic review was to assess the long-term impact of early sport specialization on injury rates during sport participation in a sample of collegiate, national, and professional athletes. Early sport specializers perform their sport-specific motor pattern excessively during their developmental stage and commonly suffer from overuse injuries[10]. Such pattern could lead to overactive muscle groups, while multisport and late sport specializers benefit from a more developed motor control, psychological, and biomechanical performance[15,37]. Thus, we hypothesized that an elite athlete who specialized in their sport from early adolescence would have a higher likelihood of suffering from a non-contact injury compared to those who delayed sport specialization. While the results indicated fewer injuries in the ESS cohort than non-ESS (OR=0.731, p-value > 0.05), the results were not statistically significant. There was a high degree of heterogeneity among studies with evidence of publication bias, which suggest that the true association between ESS and injury rates may be stronger than what was observed in the included studies. All the included studies were nonclinical trial and retrospective, which could explain the high degree of heterogeneity and publication bias in the results.

The risks and benefits surrounding ESS has been a discussion topic over the past few decades especially in the U.S., and the rates and risks of injuries during adolescence is amply reported. Although injury rates increase following an initial injury, the rates of non-contact sport injuries in collegiate or professional sports participation cannot be directly assumed from the rates of injuries in adolescents. In very recent years, however, limited peer-reviewed evidence has surfaced that evaluated early sport specialization patterns of athletes and their sport participation at collegiate, national, or professional competitions (i.e., *elite sport participation*), and to the best of our knowledge, this is the first systematic review with quantitative analysis to assess their findings.

The notion among sport professionals supports delaying most sport specialization in adolescents to reduce risks associated with ESS in most sports, especially in the absence of evidence that ESS is required to achieve success at elite levels or improve task-related performance outcomes during adolescence [8,10,14,15,17,38–40]. However, it is also necessary for an athlete to gain the sport-specific skills to excel in their sports, and some athletes may require higher coach-led exposure hours than others to reach a competitive status. Perhaps a compromise may be optimal, where for every main-sport coach-led exposure hour, there is a predetermined unstructured and free-play agenda for those adolescent athletes.

This method may serve the sport-specific skill attainment required to excel at a sport, while allowing the adolescent athlete to benefit from a nurturing and joyful environment.

### Limitation

All the included studies are retrospective, which means they were not designed to test the causal relationship between ESS and injury rates. Another limitation was that male and female athletes from the same sport were not reported in the included studies. Additionally, the search strategy allowed for reports on sports from all countries, yet, only U.S. sports teams were represented in this study due to lack of published evidence, even though some sports are widely considered more represented and competitive in other countries (e.g., football/soccer). Moreover, the heterogeneity of the study designs (e.g., survey-based or online databases were assessed, types and severity of injury were reported, recurrent versus first-time injury reports, etc.) and terminology used in this field is a limitation of this study, and there have been attempts to unify the scientific community to use a single method of reporting the findings for early sport specialization[6]. The level of expertise by athletes at the time of the injury rate report (e.g., collegiate, adult professional, adult national and Olympians, etc.) could have also an impact on the findings, but due to the limited number of studies and the sample size, a moderator analysis in this meta-analysis was not meaningful.

## Conclusion

Findings of this study suggest that there is no clear evidence that ESS increases the risk of injury in elite sport participation. It is important to note that the findings of this study do not mean that ESS is risk-free. It is still possible for athletes who specialize in a single sport to experience injuries. Our review highlights the need for further prospective research to better understand the potential impact of early sport specialization on the risks of non-contact injuries in elite sport participation. Future research should clearly define their study design and report the data for future analysis and transparency. In addition, a more equal representation of sports across different countries and genders for the same sports (e.g., men’s versus women’s basketball), where applicable, is highly encouraged.

## Data Availability

All data produced in the present study are available upon reasonable request to the authors.

## Author contributions

Bahman Adlou originated the study idea, created study design, search strategy, pooled studies, reviewed them, extracted applicable data for analysis, carried out the statistical analysis procedure(s), visualized the findings, and drafted the manuscript and confirmed the final version for submission. Dr. Wendi Weimar helped with shaping the research question and confirmed the design, carried out the search strategy and evaluated the pooled studies to aggregate potential studies to include in the systematic review and meta-analysis, was involved in the final consensus on the included data for the review, extracted the data for further analysis, and provided valuable recommendations for the final manuscript draft. Dr. Christopher Wilburn served as the tiebreaker for the included studies where applicable, was involved in the consensus on included studies, helped with data extraction where necessary, and provided valuable recommendations for the final manuscript draft. Dr. Alan Wilson helped with the research design, reviewed the statistical analysis and helped in confirming the optimal analysis and the visualization of the data, and provided valuable recommendations for the final manuscript draft. All authors read and approved this final version of the manuscript and agree with the order of presentation of the authors.

## Acknowledgment

Special thanks to Auburn University Librarians for providing guidance over the comprehensive search. The authors declare that they have no competing interests. All data generated or analyzed during this study are included in this published article. R language script used for analysis is included in the submitted supplementary materials. This research did not receive any specific grant from funding agencies in the public, commercial, or not-for-profit sectors.

